# Effect of Early Treatment Intensity on Progression Independent of Relapse Activity and Disability Accumulation in Multiple Sclerosis

**DOI:** 10.1101/2025.07.18.25331653

**Authors:** Jessica Jane Ye, Negar Molazedeh, Mariann Polgar-Turcsanyi, Tanuja Chitnis

## Abstract

**Background and Objectives:** Treatment initiation strategies for disease modifying therapy in multiple sclerosis (MS) are debated, with recent trends favoring induction over escalation approaches. Whether this impacts risk of progressive disease and long-term accumulation of disability is unclear.

**Methods:** An observational study of a real-world cohort of patients diagnosed with MS registered in the Comprehensive Longitudinal Investigation of Multiple Sclerosis at Brigham and Women’s Hospital study and the Massachusetts General Hospital Pediatric Multiple Sclerosis Database was performed. Patients 12-50 years old at diagnosis were grouped by treatment initiated within 6 months of diagnosis into low-efficacy early therapy (LEET) and high-efficacy early therapy (HEET) groups. Risks of progression independent of relapse activity (PIRA), relapse associated worsening (RAW), and sustaining an Expanded Disability Status Scale (EDSS) score ≥ 5.0 were calculated.

**Results:** 750 patients were analyzed (*n* = 583 LEET, 112 HEET). HEET vs LEET groups had a similar risk of PIRA (adjusted hazard ratio = 0.99, *P*-adjusted = 0.95) but a significantly lower risk of RAW (adjusted hazard ratio = 0.48, *P*-adjusted = 0.015). Sub-stratification of the HEET group into high- and very high-efficacy early therapy groups (i.e. ocrelizumab, natalizumab) showed similar risk of PIRA. Cox proportional hazards analysis revealed older age at diagnosis as a significant contributor to risk of PIRA. Nonetheless young patients still had substantial risk of PIRA, which was similar regardless of treatment efficacy group. In a subset of patients with MRI data, lower brain parenchymal also significantly increased risk of PIRA. LEET, HEET, and VHEET groups also had a similar risk of sustaining an EDSS score ≥ 5.0. HEET and VHEET patients, however, had a slightly lower change in EDSS over time.

**Discussion:** While HEET reduces risk of RAW, it does not significantly affect the risk of PIRA, even in patients diagnosed before age 30. It also does not significantly affect the risk of sustaining an EDSS score ≥ 5.0, though may minimally decrease change in EDSS/year. This suggests inductive versus escalation strategies primarily influence patient outcomes by decreasing RAW, and underscores the need for emerging therapeutics to target PIRA, which continues to cause significant disability in patients on treatment.

## Introduction

Since the proliferation of different types of disease modifying therapies (DMTs) in multiple sclerosis (MS), there has been debate about the best initiation strategies to maximize benefit and minimize risk.^1,2^ In a more traditional escalation approach, lower-efficacy, lower-risk immunosuppressive agents are started first and switched to more aggressive immunotherapies after breakthrough disease is observed. In contrast, in an early-intensive induction approach, high-efficacy immunotherapy is initiated first to quickly quell disease, then de-escalated as disease activity wanes. Increased safety of, familiarity with, and expanded access to high-efficacy therapies, as well as studies showing decreased relapse rate and lower average increase in disability scores,^3–7^ have fueled recent trends toward starting high-efficacy therapies early. However, the mechanism(s) by which early use of stronger immunosuppressing therapies has an impact on long-term disability accumulation, especially in the setting of progressive disease, is not clear. Observational studies have demonstrated poor correlation between relapses and silent progression.^8,9^ Ongoing prospective randomized clinical trials including TREAT-MS and DELIVER-MS aim to further assess the relative benefits in early initiation of high-efficacy therapies.^10–12^

A measurement concept recently defined to capture silent progression in MS is progression independent of relapse activity (PIRA), as distinct from relapse-associated worsening (RAW).^13–16^ PIRA and RAW are two types of confirmed disability accumulation (CDA) events, calculated via changes in clinical functional scores like the Expanded Disability Status Scale (EDSS), and categorized by time proximity to clinical relapses. Though relapses have long been the focus of MS treatment, recent studies show PIRA drives the bulk of disability accumulation in MS patients, even in relapsing-remitting subtypes.^13,17^ PIRA has been described to occur within years of disease onset, in both young and older MS patients.^18,19^ Its occurrence is also associated with increased disability accumulation long term,^20^ and thought of as a manifestation of smoldering disease.^21^

As such, PIRA is a metric of silent progression in MS which may be sub-optimally controlled by modern DMTs. The objective of our study is to examine PIRA, contextualized with RAW and change in EDSS, to assess whether treatment initiation strategy with low-efficacy early therapy (LEET) versus high-efficacy early therapy (HEET) affects silent progression in MS.

## Materials and methods

### Data

A retrospective, real-world cohort was assembled from patients enrolled in the Comprehensive Longitudinal Investigation of Multiple Sclerosis at Brigham and Women’s Hospital (CLIMB) study in the Harvard Multiple Sclerosis Patient Data Base and the Massachusetts General Hospital Pediatric Multiple Sclerosis Database from January 1, 1999, to December 1, 2023 who met the following inclusion criteria: (1) diagnosis of MS by 2017 McDonald Criteria,^22^ (2) age at diagnosis 12-50 years old, (3) started on DMT within 6 months of diagnosis, (4) maintained on DMT for at least 5 years, and (5) EDSS 0-4.5 at screening visit. EDSS scores were entered by MS neurologists trained in Neurostatus EDSS conduct. Patients were seen in the context of routine clinical care at Brigham and Women’s Hospital or Massachusetts General Hospital, with regular follow up typically every 6 months.

Demographic data, MS subtype (relapsing remitting (RR), secondary progressive (SP), primary progressive (PP), or progressive relapsing (PR)) as of last visit, treatment data including dates of treatment, flare/attack dates, and visit dates with associated visit EDSS were retrieved for each patient via an Oracle database. Baseline MRI measurements were also retrieved where recorded. The confidentiality of the data is maintained according to the Mass General Brigham institutional policy. This study is approved by the Mass General Brigham Institutional Review Board.

### Treatment Group Definitions

Patients were grouped into low efficacy (LE) early treatment (LEET) and high-efficacy (HE) early treatment (HEET) groups based on the first treatment type they started within 360 days of diagnosis and continued for ≥165 days (∼5.5 months) (**Supplemental Figure 1**). This was designed to best capture a patient’s primary initial treatment in cases where there was a rapid change in therapy type or discontinuation of treatment (assuming that a disease modifying therapy takes 5-6 months to take full effect) and differentiate this from a true treatment change. For instance, if a patient received natalizumab for 1 month, then switched to glatimer acetate for the next 6 months, the patient was categorized as LEET. But if a patient received natalizumab for ≥165 days and switched to glatimer acetate afterward, it was counted as a de-escalation and the patient was categorized as HEET. If no treatment in the first 360 days post-diagnosis lasted ≥165 days, then the patient was sorted as ‘unclear’ (which captures patients with unknown treatment at diagnosis or poorly sustained treatment). All patients in the dataset started therapy within 6 months of diagnosis given the inclusion criteria. To look with further resolution at the effect of moderately high or very high efficacy treatments on disability accumulation, we further stratified the original HEET group into high efficacy (HE) and very high efficacy (VHE) groups (HEET, VHEET). Demographics and initial EDSS of the patients in each category are summarized in **Table 1**. Designation of specific therapies as LE, HE, or VHE are summarized in **Figure 1**.

**Figure 1.**
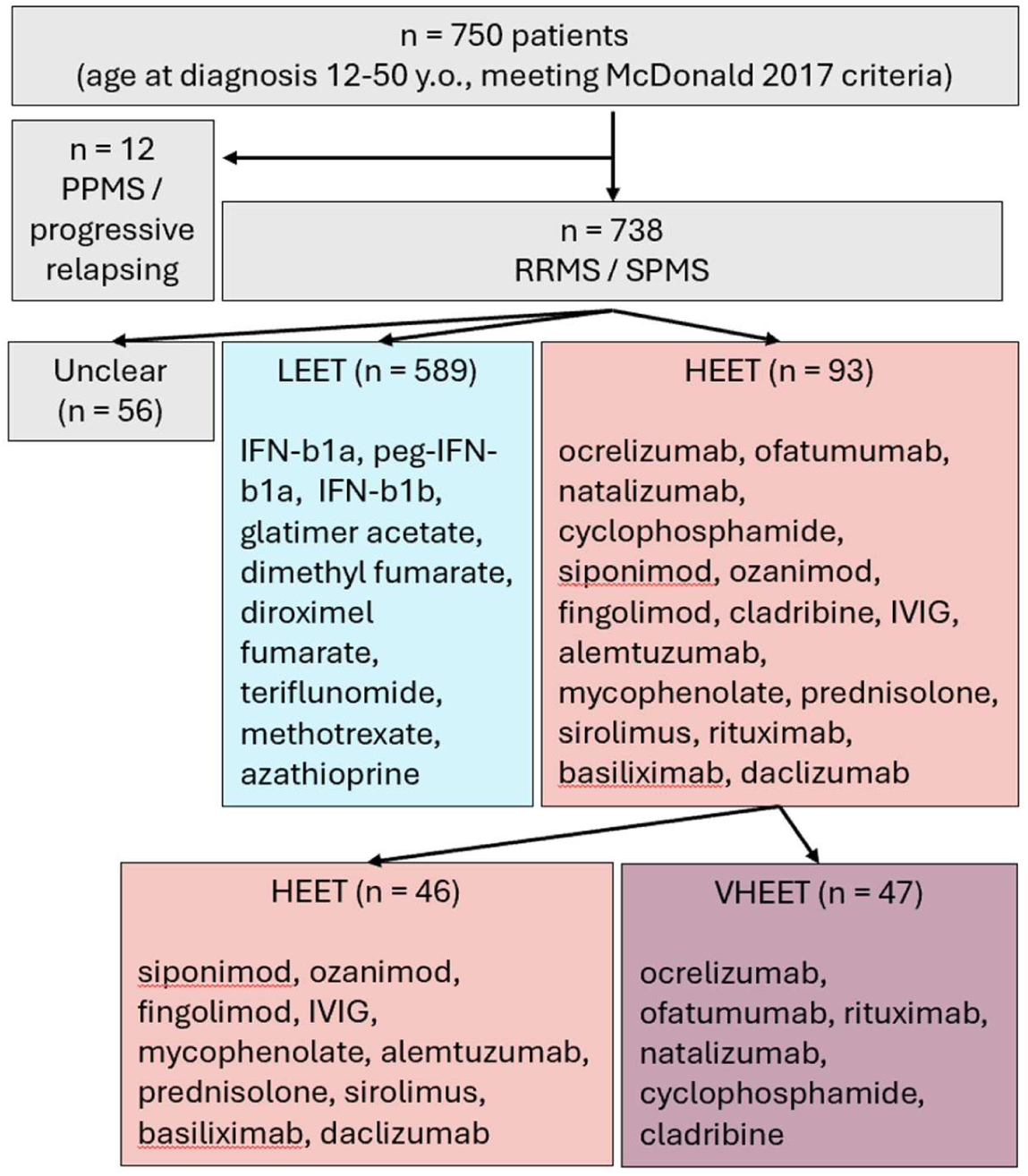
Flow diagram of patients in the study cohort and analysis groups. From the base cohort, analysis was focused on patients with RRMS and SPMS. Sorting into treatment groups by early treatment type was performed as described for both the general cohort and the RRMS/SPMS cohort. For simplicity, only numbers for the RRMS/SPMS cohort are displayed here (see **Table 1** for additional details). Specific medications grouped as HE and LE therapies are listed, as are the HE therapies sub-stratified as VHE therapies.

**Table 1.** Cohort Patient Demographics by Treatment Groups.

|  | **Total**<br>**(N=750)** | **Unclear**<br>**(N=57)** | **LEET**<br>**(N=596)** | **HEET**<br>**(N=97)** | **HEET**<br>**(N=47)** | **VHEET**<br>**(N=50)** |
| --- | --- | --- | --- | --- | --- | --- |
| **Sex** |  |  |  |  |  |  |
| Male | 196 (26.1%) | 9 (15.8%) | 153 (25.7%) | 34 (35.1%) | 19 (40.4%) | 15 (30.0%) |
| Female | 554 (73.9%) | 48 (84.2%) | 443 (74.3%) | 63 (64.9%) | 28 (59.6%) | 35 (70.0%) |
| **Age at diagnosis (years)** |  |  |  |  |  |  |
| Mean (SD) | 33.0 (9.53) | 29.6 (9.91) | 33.3 (9.42) | 33.5 (9.66) | 33.9 (10.7) | 33.1 (8.69) |
| **Race/ethnicity** |  |  |  |  |  |  |
| American Indian, Alaska Native | 3 (0.4%) | 0 (0%) | 3 (0.5%) | 0 (0%) | 0 (0%) | 0 (0%) |
| Asian | 5 (0.7%) | 0 (0%) | 4 (0.7%) | 1 (1.0%) | 1 (2.1%) | 0 (0%) |
| Black or African American | 45 (6.0%) | 3 (5.3%) | 39 (6.5%) | 3 (3.1%) | 1 (2.1%) | 2 (4.0%) |
| East Asian, South-East | 2 (0.3%) | 1 (1.8%) | 1 (0.2%) | 0 (0%) | 0 (0%) | 0 (0%) |
| More than one race | 27 (3.6%) | 0 (0%) | 22 (3.7%) | 5 (5.2%) | 4 (8.5%) | 1 (2.0%) |
| Native Hawaiian, Pacific Islander | 1 (0.1%) | 0 (0%) | 1 (0.2%) | 0 (0%) | 0 (0%) | 0 (0%) |
| South Asian | 4 (0.5%) | 0 (0%) | 4 (0.7%) | 0 (0%) | 0 (0%) | 0 (0%) |
| Unknown or not reported | 16 (2.1%) | 3 (5.3%) | 10 (1.7%) | 3 (3.1%) | 2 (4.3%) | 1 (2.0%) |
| White | 647 (86.3%) | 50 (87.7%) | 512 (85.9%) | 85 (87.6%) | 39 (83.0%) | 46 (92.0%) |
| **EDSS at first visit** |  |  |  |  |  |  |
| Mean (SD) | 1.39 (1.02) | 1.61 (1.21) | 1.32 (0.996) | 1.68 (0.974) | 1.60 (0.895) | 1.76 (1.05) |
| **time fraction on HE DMT** |  |  |  |  |  |  |
| Mean (SD) | 0.318<br>(0.345) | 0.268<br>(0.325) | 0.242<br>(0.291) | 0.814<br>(0.237) | 0.851<br>(0.208) | 0.779<br>(0.258) |
| **time fraction on LE DMT** |  |  |  |  |  |  |
| Mean (SD) | 0.520<br>(0.361) | 0.309<br>(0.343) | 0.617<br>(0.316) | 0.0456<br>(0.129) | 0.0280<br>(0.0631) | 0.0621<br>(0.168) |
| **time observed (years)** |  |  |  |  |  |  |
| Mean (SD) | 10.7 (5.83) | 7.65 (5.63) | 11.7 (5.68) | 6.74 (4.38) | 7.01 (3.72) | 6.48 (4.95) |
| **MS phenotype** |  |  |  |  |  |  |
| Primary Progressive | 9 (1.2%) | 1 (1.8%) | 4 (0.7%) | 4 (4.1%) | 1 (2.1%) | 3 (6.0%) |
| Progressive Relapsing | 3 (0.4%) | 0 (0%) | 3 (0.5%) | 0 (0%) | 0 (0%) | 0 (0%) |
| Relapsing-Remitting | 664 (88.5%) | 55 (96.5%) | 520 (87.2%) | 89 (91.8%) | 44 (93.6%) | 45 (90.0%) |
| Secondary Progressive | 74 (9.9%) | 1 (1.8%) | 69 (11.6%) | 4 (4.1%) | 2 (4.3%) | 2 (4.0%) |
| **1.5T baseline T2 volume** |  |  |  |  |  |  |
| Mean value (SD) | 3.46 (2.62) | 2.32 (1.80) | 3.58 (2.67) | 2.99 (2.81) | 2.99 (2.81) | NA (NA) |
| Entries missing data (%) | 675 (90.0%) | 52 (91.2%) | 530 (88.9%) | 93 (95.9%) | 43 (91.5%) | 50 (100%) |
| **1.5T baseline BPF (%)** |  |  |  |  |  |  |
| Mean value (SD) | 88.3 (3.12) | 89.4 (2.21) | 88.3 (3.20) | 86.8 (2.66) | 86.8 (2.66) | NA (NA) |
| Entries missing data (%) | 675 (90.0%) | 52 (91.2%) | 530 (88.9%) | 93 (95.9%) | 43 (91.5%) | 50 (100%) |
| **3T baseline T2 volume** |  |  |  |  |  |  |
| Mean value (SD) | 4.43 (6.94) | 2.63 (NA) | 4.09 (5.93) | 5.45 (9.83) | 1.78 (0.759) | 9.12 (14.2) |
| Entries missing data (%) | 730 (97.3%) | 56 (98.2%) | 583 (97.8%) | 91 (93.8%) | 44 (93.6%) | 47 (94.0%) |
| **3T baseline BPF (%)** |  |  |  |  |  |  |
| Mean value (SD) | 85.9 (2.46) | 86.1 (NA) | 85.9 (2.65) | 85.8 (2.48) | 86.8 (3.25) | 84.9 (1.45) |
| Entries missing data (%) | 730 (97.3%) | 56 (98.2%) | 583 (97.8%) | 91 (93.8%) | 44 (93.6%) | 47 (94.0%) |

Legend: DMT = disease modifying therapy, N = number of patients, SD = standard deviation. Demographics for sub-stratification of HEET to HEET and VHEET displayed at right.

### Calculation of Approximate Adherence

Adherence to HE or LE therapy was approximated by calculating the number of days each patient was actively on each type of therapy based on start and end dates, and dividing this by the total number of days in observation in the database (up to a maximum of 20 years). This was represented as the LE or HE time fraction (**Table 1**). LE and HE time fractions were calculated for patients in each therapy group.

### Definition of PIRA and RAW events

Using EDSS and clinical attack data gathered longitudinally throughout sequential clinic visits, PIRA and RAW events were calculated based on 2025 consensus guidelines^16^ using a roving baseline approach.^15,16,23^ Briefly, disability accrual was defined as an increase from the reference EDSS, scaled by reference EDSS in the following manner (+1.5 for EDSS 0-1, +1.0 for EDSS 1-5.5, +0.5 for EDSS ≥6) to account for non-linearity in the EDSS. Disability accrual was confirmed if the EDSS at a following visit ≥12 months after the incident visit met the same increase criteria as well as all intervening visits. A CDA event was called PIRA if there were no relapses between the index visit and the visit prior, and no relapses in the 30 days before each confirmation score. If there was an increase on the EDSS that met the disability accrual definition with a relapse since the previous visit, this was called a RAW event. Baseline EDSS scores were initialized using the first visit score, but re-baselined after confirmed PIRA, RAW, or confirmed disability improvement (CDI) events (scaled *decreases* in EDSS score sustained over a follow up visit ≥180 days later). After PIRA or CDI events, the new baseline was set to the EDSS at the confirmation visit. After a RAW event, the new baseline EDSS was set to the EDSS at a clinical visit >30 days after the relapse.

### Calculation of baseline MRI T2 lesion volume and brain parenchymal fraction

Brain parenchymal fraction (BPF) and T2 lesion volume (T2LV) were calculated from dual-echo, conventional spin-echo MRI images (DE-CSE) acquired on 1.5 T or 3T Signa GE scanners at BWH using a standard quadrature head coil or multichannel head coil (8 Channel High Resolution Brain Array (8HR BRAIN)) as described previously.^24,25^ In brief, for 1.5T DE-CSE MRI protocols were acquired axially with pulse sequences including various combinations of following parameter ranges: TR = 2216–3000 ms, TE1/TE2 30/80 ms, slice thickness 3 mm, with no interslice gaps, resulting in a pixel size of 0.7813–0.9375 mm. Quantitative image analysis was performed from the DE-CSE images using an automated template-driven segmentation (TDS) pipeline with partial volume effect correction to derive the intracranial volume (ICV), gray matter (GM), CSF, white matter (WM), and white matter lesions.^26^ BPF was calculated by the following formula: BPF = (GM + WM + lesions)/ICC,^27^ where ICC is the volume of the intracranial cavity serving as reference for individual head size.^28^ This was followed by manual editing of output segmentation maps by an experienced observer using 3D Slicer software.^29^ We note that no correction for misclassification of T1 hypointensities was performed in our pipeline, but recent work from our group has demonstrated that these hypointensities have a limited impact on measures of BPF.^30^ For 3T images, T1, T2, and fluid-attenuated inversion-recovery (FLAIR) images were acquired sagittally with 1 mm isotropic voxel size and analyzed using 3T morphometry.^25^

### Calculation of delta EDSS and dEDSS/dt

For the observation period (time to censorship or 20 years), the overall change in EDSS score from first to last visit was defined as the ΔEDSS. This was normalized to the period of observation or time to censorship to calculate dEDSS/dt (change in EDSS per year).

### Statistical Analysis

Statistical analysis was performed in R using the following packages. Survival analysis and Cox proportional hazards were performed with the standard functions in the *survival* and *survminer* library. Adjusted *P*-values considered patient age at diagnosis, gender, race, and first visit EDSS as potentially confounding variables in addition to treatment group. In the subset of patients w MRI data, baseline MRI T2 lesion volume and brain parenchymal fraction were also included as confounding variables. *P*-values displayed on the survival curve plot represent a log rank test *P*-value. Adjusted survival curves obtained using the ggadjustedcurves function after fitting Cox proportional hazards model. To compare dEDSS/dt by treatment group, analysis of variance in the standard *stats* library, followed by Tukey’s post-hoc analysis using the general linear hypothesis testing *glht* function in the *multcomp* library, was used to obtain unadjusted *P*-values. To adjust for additional covariates as above, the *lm* function was used. For these calculations, patient race was simplified into “white” and “non-white” to avoid infinite values caused by small sample numbers.

### Standard Protocol Approvals, Registrations, and Patient Consents

Institutional Review Board approval was granted by the Mass General Brigham Human Research Protection Program for the collection and analysis of the data in this study.

### Data availability

Deidentified data allowing for independent verification of research results will be made available upon reasonable request from a qualified investigator in a manner compliant with prior patient consent and institutional policy for patient privacy and protection of personal health information.

## Results

### Cohort

750 patients aged 12-50 years at time of diagnosis with MS by McDonald Criteria who had recorded longitudinal treatment and EDSS data were analyzed in this retrospective observational study. Demographic data for these patients are summarized in **Table 1**. Groupings of the patients into ‘unclear,’ LEET, and HEET groups were established based on initial treatment started within 6 months of diagnosis sustained for ≥165 days (see Methods). For additional analysis of treatment type within the HEET group, we further stratified the HEET group into patients receiving very high efficacy early treatment (VHEET) like anti-CD20 therapies, natalizumab, cladribine, and cyclophosphamide, from other high efficacy treatments (S1P modulators, mycophenolate, sirolimus, etc.) (**Figure 1**). Demographics of the sub-stratified patients are shown to the right of **Table 1**. Overall patient characteristics including age at diagnosis, gender, ethnicity, and MS phenotype were similar in each group except for EDSS at initial visit, which was lower in LEET vs HEET/VHEET groups (**Supplemental Table 1**). This may reflect inclinations to use step-up therapy for patients presenting with less disability, and early high intensity therapy for those presenting with more symptoms. Most patients (>85%) in all groups had RRMS, which reflects the population seen at our center. Most patients are also of white race/ethnicity in all groups, reflecting typical patterns of involvement in research. Additionally, many more patients were in the LEET vs. HEET group, likely as HE therapies were not available in the earlier years of the database.

To broadly assess how much initial treatment type was sustained throughout the treatment course, the fraction of time in observation on each type of treatment (LE/HE fraction) was calculated. Patients in the HEET and VHEET groups in general had a higher adherence to HE/VHE therapy (∼80% of time in observation on average), whereas those in the LEET group stayed on LE therapy for ∼60% of time in observation.

A subset of patients had baseline MRI measurements recorded in the database (74 1.5T, 30 3T MRIs). Due to differences in technique, measurements derived from 1.5T MRIs were kept separate from those from 3T MRIs. Overall, in this subset, no significant differences were observed in the baseline MRI measures of patients started on HEET vs. LEET (**Supplemental Table 1**).

### Intensity of initial therapy does not affect risk of PIRA but lowers risk of RAW

To determine whether intensity of early treatment affects disability accumulation, we first assessed its impact on risk of PIRA or RAW as defined by 2025 consensus guidelines^16^. We also performed the analysis using 2023 consensus guidelines with a 180 day confirmation period and −90/+30-day attack window^15^ and did not observe any significant differences. We based PIRA/RAW calculations only on EDSS score as it was the most available metric and encapsulates a high-level view of patient disability. Event-free survival was calculated based on time to first event. To focus our comparisons, we limited our primary analysis to patients in a defined early therapy group (no “unclear” group) diagnosed with either RRMS or SPMS, as they are felt to have similar MS biology. Interestingly, this analysis revealed no significant difference in the relative risk of PIRA over time between patients in HEET vs LEET groups (**Figure 2A**, **Table 2**). However, there was a significantly lower risk of RAW in the HEET group (**Figure 2B**, **Table 2**). These trends were sustained after adjusting for potentially confounding variables (age at diagnosis, gender, ethnicity, and EDSS at first visit) using a Cox proportional hazards model. Including all patients with “unclear” early treatment and other MS subtypes also did not significantly change analysis results (**Supplemental Figure 2A-B**).

**Figure 2.**
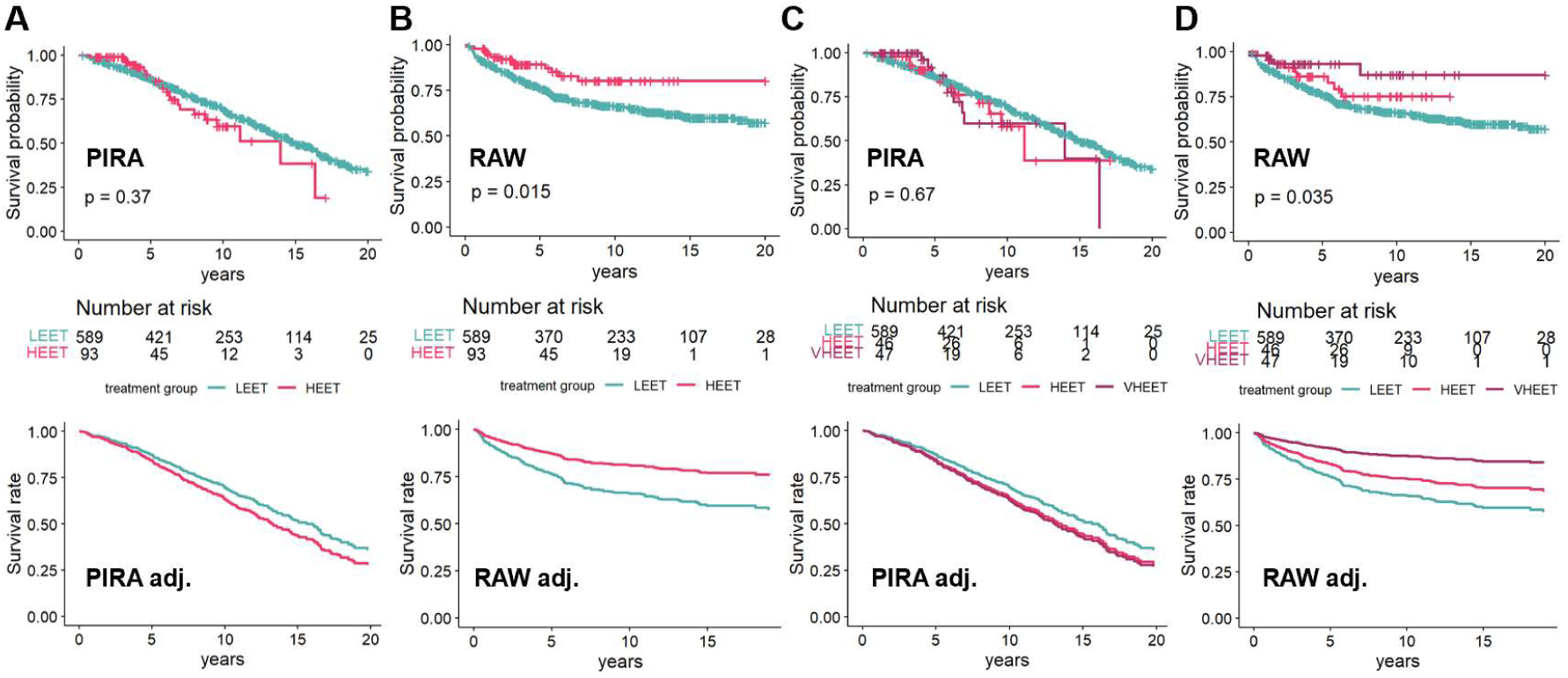
Higher efficacy early treatment significantly lowers risk of RAW but does not significantly affect risk of PIRA. Graphs of event-free survival over time. Unadjusted survival curves with risk table displayed over adjusted survival curves after fitting a Cox proportional hazards model. P-values displayed are the result of log-rank tests; see **Table 2** for hazard ratios. **(A, B)** Survival to first PIRA **(A)** or RAW **(B)** event in patients with HEET vs LEET. **(C, D)** Survival to first PIRA **(C)** or RAW **(D)** event in patients with LE, HE, or VHE early treatment.

**Table 2.** Calculation of Hazard Ratios and Mean Differences Across Treatment Groups.

| Outcome | Unadjusted |  | Adjusted |  |
| --- | --- | --- | --- | --- |
| **PIRA\*** |  |  |  |  |
| LEET | Reference |  | Reference |  |
| HEET | HR = 1.23 (95% CI = 0.79-1.91, p = 0.37) |  | HR = 1.26 (95% CI = 0.81-1.98, p-adj = 0.31) |  |
| LEET | Reference |  | Reference |  |
| HEET | HR = 1.24 (95% CI = 0.69-2.24, p = 0.47) |  | HR = 1.24 (95% CI = 0.69-2.24, p-adj = 0.48) |  |
| VHEET | HR = 1.20 (95% CI = 0.64-2.28, p = 0.57) |  | HR = 1.30 (95% CI = 0.68-2.47, p-adj = 0.43) |  |
| **RAW\*** |  |  |  |  |
| LEET | Reference |  | Reference |  |
| HEET | HR = 0.51 (95% CI = 0.29-0.88, p = 0.017) | * | HR = 0.51 (95% CI = 0.29-0.89, p-adj = 0.018) | * |
| LEET | Reference |  | Reference |  |
| HEET | HR = 0.66 (95% CI = 0.34-1.29, p = 0.22) |  | HR = 0.68 (95% CI = 0.35-1.33, p-adj = 0.26) |  |
| VHEET | HR = 0.33 (95% CI = 0.12-0.88, p = 0.027) | * | HR = 0.32 (95% CI = 0.12-0.86, p-adj = 0.024) | * |
| **susEDSS ≥5\*** |  |  |  |  |
| LEET | Reference |  | Reference |  |
| HEET | HR = 1.21 (95% CI = 0.48-3.05, p = 0.70) |  | HR = 0.91 (95% CI = 0.36-2.33, p-adj = 0.85) |  |
| LEET | Reference |  | Reference |  |
| HEET | HR = 1.43 (95% CI = 0.44-4.63, p = 0.55) |  | HR = 1.15 (95% CI = 0.35-3.78, p-adj = 0.82) |  |
| VHEET | HR = 0.98 (95% CI = 0.24-4.03, p = 0.98) |  | HR = 0.70 (95% CI = 0.17-2.92, p-adj = 0.62) |  |
| **dEDSS/dt\*** |  |  |  |  |
| LEET | Reference |  | Reference |  |
| HEET | Mean difference = -0.11 (s.e. = 0.031, p = 0.0004) | *** | Mean difference = -0.081 (s.e. = 0.029, p-adj = 0.006) | ** |
| LEET | Reference |  | Reference |  |
| HEET | Mean difference = -0.075 (s.e. = 0.042, p = 0.17) |  | Mean difference = -0.055 (s.e. = 0.040, p-adj = 0.17) |  |
| VHEET | Mean difference = -0.15 (s.e. = 0.042, p = 0.001) | ** | Mean difference = -0.11 (s.e. = 0.040, p-adj = 0.007) | ** |

Legend: \* Results use Cox proportional hazards regression. + Results use linear regression. Adjusted analysis accounts for demographic covariates of age at diagnosis, race (white or non-white), and initial EDSS at first visit; unadjusted analysis only compares the treatment groups. Analysis only including RRMS/SPMS patients assigned an early treatment group (no 'unclear' group). Units of dEDSS/dt are EDSS points/year.
Significance codes: 0 '\*\*\*' 0.001 '\*\*' 0.01 '\*' 0.05 '.' 0

To assess whether these observations were due to inclusion of more moderate efficacy treatments in the HEET group, we further stratified the HEET group into those receiving VHE treatment (i.e. ocrelizumab and natalizumab) and other HE treatment (**Figure 1**). However, patients on VHEET still did not have a significantly different risk of PIRA than those on HEET or LEET (**Figure 2C**, **Table 2**). The risk of RAW was again significantly lower in the VHEET vs LEET group in adjusted and unadjusted analyses, while HEET had an intermediate effect (**Figure 2D**, **Table 2**).

### Age at diagnosis and baseline brain parenchymal fraction affect risk of PIRA

Given that treatment group did not affect risk of PIRA over time, we investigated what other factors might play a role. Cox proportional hazards analysis of all covariates affecting survival time to first PIRA showed age at diagnosis was significantly associated with risk of PIRA (**Supplemental Table 2**). Testing age ranges 12-20 years, 20-30 years, 30-40 years, and 40-50 years at diagnosis, it was found that a transition point occurred around age 30, where those patients diagnosed after 30 years of age had a higher risk of PIRA than those diagnosed before age 30 years, which had intermediate and low risk (**Figure 3A**). However, even among patients diagnosed before 30 years of age, almost half experienced a PIRA event within 15-20 years. To then assess whether early treatment intensity affected risk of PIRA in young patients diagnosed before 30 years of age, PIRA-free survival in this subgroup was compared between LEET and HEET groups. Similar to the main cohort, intensity of early treatment again did not affect risk of PIRA in young patients diagnosed before 30 years of age, though it still decreased risk of RAW (**Figure 3B-C**, **Table 3**, **Supplemental Figure 3**).

**Figure 3.**
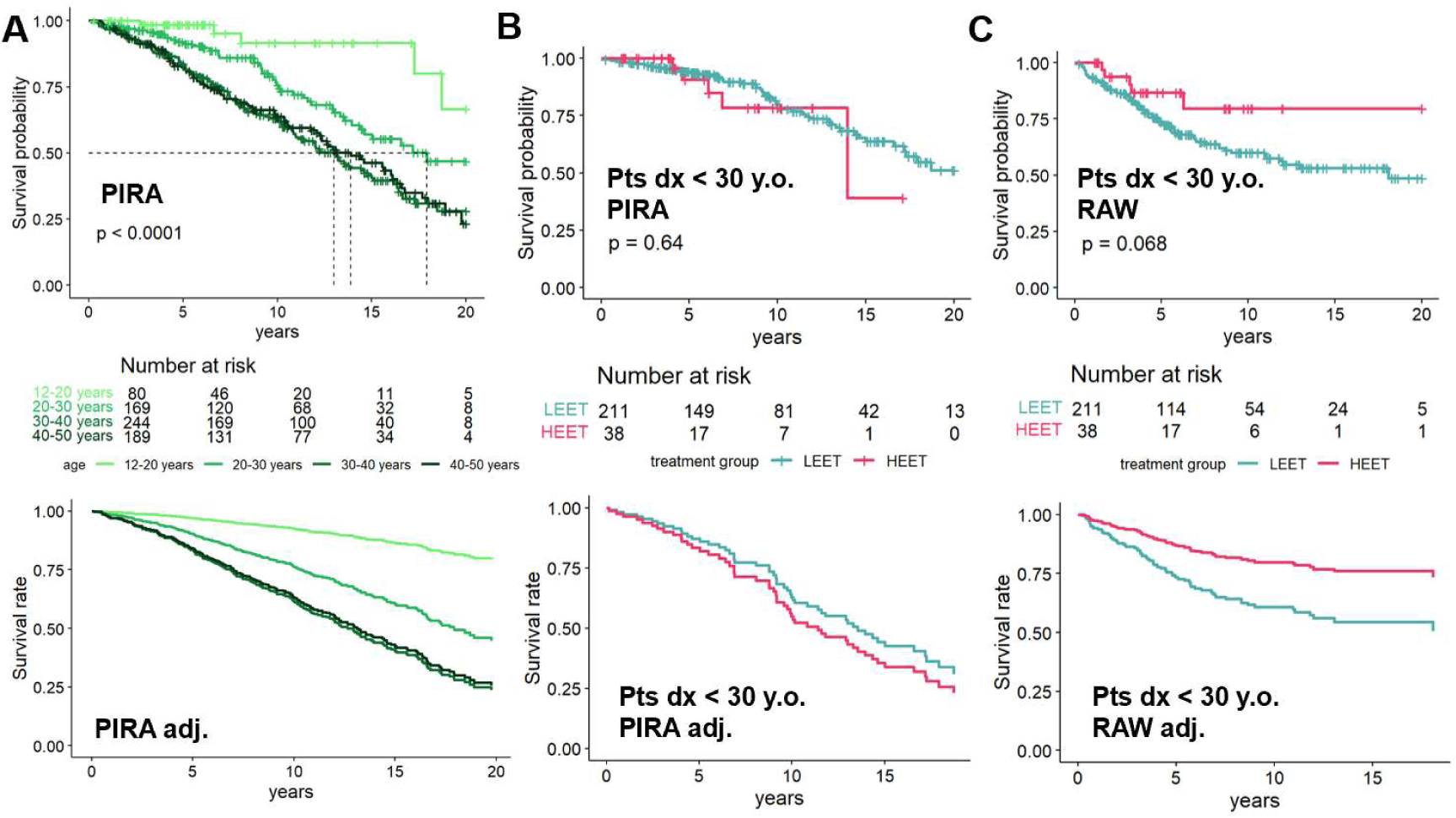
Increased age at diagnosis increases risk of PIRA. PIRA-free survival over time of RRMS/SPMS patients in a defined early treatment group. Unadjusted survival curves with risk table displayed over adjusted survival curves after fitting a Cox proportional hazards model. **(A)** PIRA-free survival in patients across all treatment groups stratified by age at diagnosis (12-20 years, 20-30 years, 30-40 years, and 40-50 years). **(B)** PIRA-free survival in young patients (diagnosed before age 30 years) with LEET or HEET. P-values displayed are the result of log-rank tests.

**Table 3.**
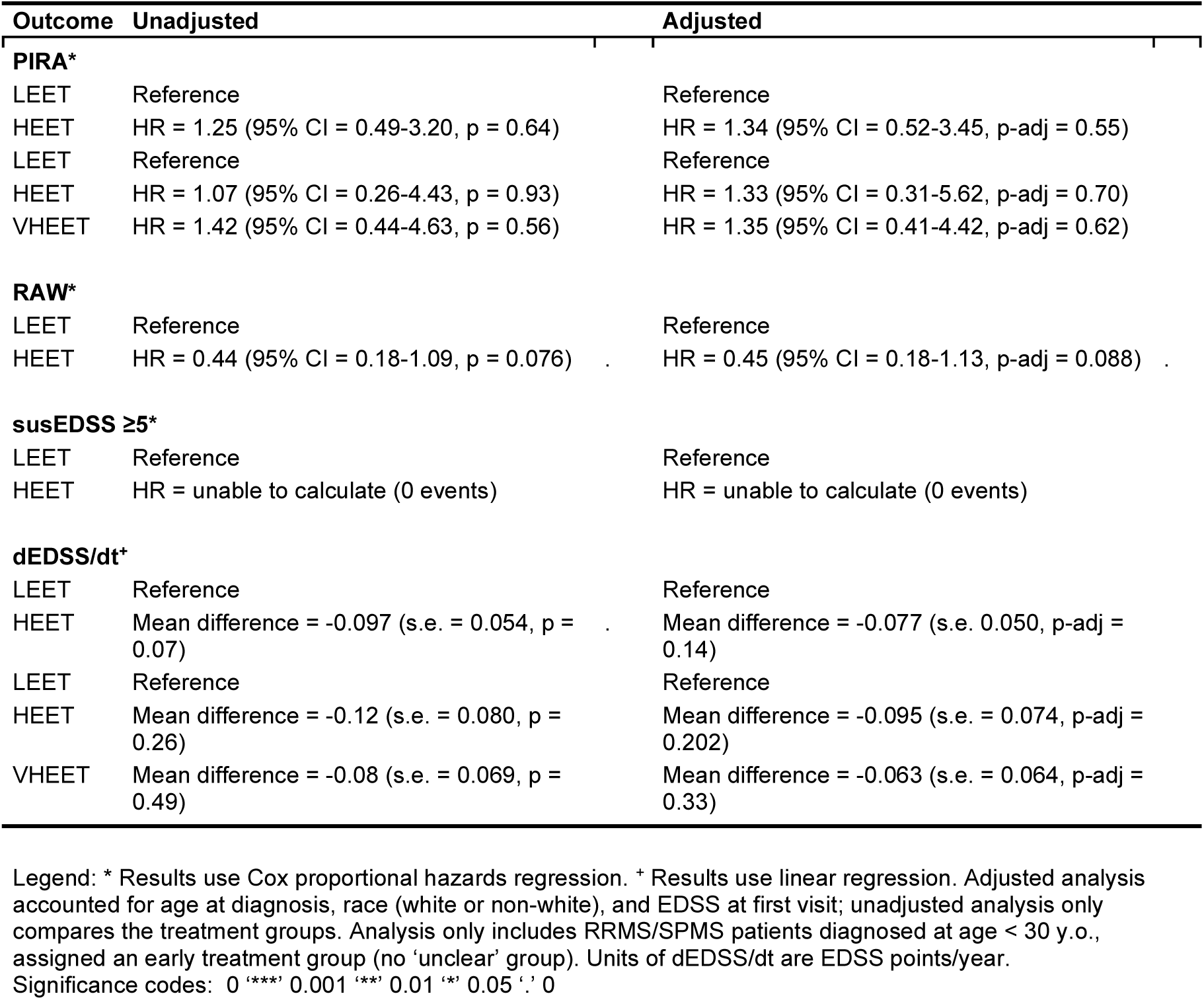
Calculation of Hazard Ratios and Mean Differences Across Treatment Groups in Young Patients (Diagnosed Before Age 30)

To assess whether baseline MRI features additionally affect risk of PIRA or RAW, a Cox proportional hazards analysis was performed on a subset of cohort patients with recorded baseline MRI measurements (**Table 1**). 69 patients with RRMS/SPMS and defined initial treatment group had baseline 1.5T MRI data, and 19 had baseline 3T MRI data. Very few PIRA or RAW events were noted in the 19 patients with 3T MRI data, so analysis was focused on those with 1.5T baseline MRI data. In this subgroup, lower brain parenchymal fraction was found to contribute significantly to a higher risk of PIRA, but not to risk of RAW (**Supplemental Table 3**). Of note, however, patients in this subgroup were almost exclusively in the LEET group as they were diagnosed before 2015 before many HE therapies were available, rendering the treatment group comparison essentially irrelevant. Thus, the contribution of baseline MRI parameters to the effect of HEET vs LEET on risk of PIRA or RAW could not be fully assessed.

### Treatment with HEET has similar risk of long-term disability as LEET, but slightly lower change in EDSS over time

To ensure capture of true disability accumulation over time, we corroborated our findings on risk of PIRA and RAW with changes in EDSS over time. First, we calculated survival time to the first visit where the patient’s EDSS reaches and becomes sustained ≥ 5.0 (where disability affects full daily activities). Consistent with the findings in PIRA, there was no significant difference in survival time to sustained EDSS ≥ 5.0 between patients started on LEET vs HEET therapy (or even VHEET therapy) (**Figure 4A-B**, **Table 2**). Cox proportional hazards analysis revealed again a significant impact of age at diagnosis, as well as an expected large impact of initial EDSS (**Supplemental Table 2**); baseline MRI parameters did not contribute. Adjusted survival curves and hazard ratios again revealed similar results.

**Figure 4.**
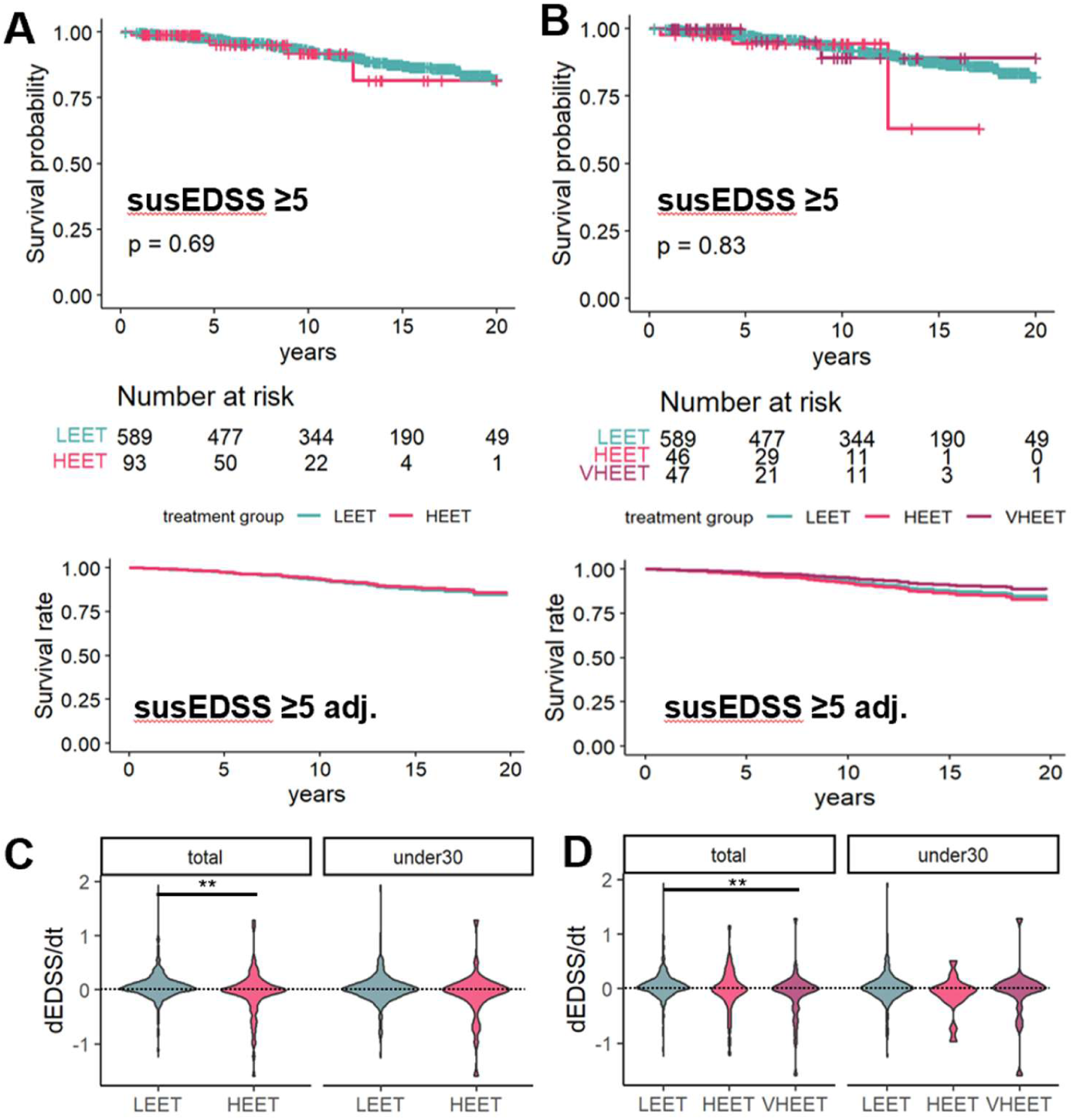
Higher efficacy early treatment does not significantly affect the risk of achieving a sustained EDSS ≥ 5.0, but slightly lowers change in EDSS over time. **(A-B)** Survival to sustained EDSS ≥ 5.0 in patients initiated LEET vs HEET **(A)** or LEET vs HEET vs VHEET **(B)**. Unadjusted survival curves with risk table displayed over adjusted survival curves after fitting a Cox proportional hazards model. P-values displayed are the result of log-rank tests; hazard ratios listed in **Table 2**. **(C-D)** Change in EDSS over time (dEDSS/dt) over the period of observation in patients initiated on LE vs HE treatment **(C)**, or LE vs HE vs VHE treatment **(D)**. Results for the primary analysis cohort (total) and young patients (under30) displayed on same axis. Significance calculated using a linear model incorporating demographic covariates; adjusted p-values displayed according to significance code in **Table 2-3**.

Because change in EDSS is heterogenous, we additionally calculated the overall change in EDSS score per year in observation (dEDSS/dt) for each patient to assess whether all or a subset of patients were contributing to the risk of progressing to sustained EDSS ≥ 5.0. Mean dEDSS/dt was slightly but significantly lower in patients on HEET versus LEET by about 0.1 EDSS points/year (**Figure 4C-D**, **Table 2**). With additional sub-stratification of HEET to HEET and VHEET, the significance of the difference was decreased due to multiple comparisons, but dEDSS/dt was still significantly lower in the VHEET group compared to the LEET group (HEET vs LEET not significant), even after adjusting for significant confounders of age and initial EDSS. Of note, change in EDSS encapsulates change from both PIRA and RAW. Thus, while there is no significant difference in risk of PIRA or achieving sustained EDSS ≥ 5.0 in patients started on LEET, HEET, or VHEET, use of HEET/VHEET may slightly slow disability accumulation, potentially by affecting risk of RAW.

As age significantly impacted risk of sustaining an EDSS ≥ 5.0, we further assessed time to sustained EDSS ≥ 5.0 in young patients diagnosed before age 30 years. Young patients overall had a lower risk of sustaining an EDSS ≥ 5.0, and no events were observed in those on HEET (preventing calculation of hazard ratio or adjusted survival curve). Despite this, however, the dEDSS/dt for young patients was similar to the general cohort, with similar breakdown by early therapy group (**Figure 4C-D**, **Table 3**).

## Discussion

In this study, we found that early treatment with high vs. low efficacy MS DMT does not significantly affect risk of PIRA in patients with RRMS/SPMS but does significantly lower risk of RAW. A sensitivity analysis isolating a subset of patients initiated on VHEET from others on HEET additionally did not show a significant effect of VHE early therapy on risk of PIRA. Though our primary analysis focused on RRMS and SPMS patients, including all MS phenotypes did not change the results. Separately examining young patients diagnosed before the age of 30, who have a lower risk of PIRA overall and theoretically less confounding of aging with PIRA, also did not show a significant impact of early treatment intensity on PIRA. HE/LE early therapy furthermore did not significantly impact a patient’s risk of sustaining an EDSS ≥ 5.0, though patients on HEET, especially VHEET, had a slightly lower average change in EDSS/year. As dEDSS/dt reflects both PIRA and RAW related changes, the difference in dEDSS/dt may predominantly reflect the decrease in RAW on HEET, as there was no significant difference in risk of PIRA on HEET. HEET may also more strongly benefit a subgroup of patients, as illustrated by the spread in the dEDSS/dt graphs, without affecting risk of PIRA or sustained EDSS ≥ 5.0 in the whole population.

These data overall imply that early HE therapy may have a limited impact on risk of PIRA, possibly because it does not adequately target the pathological mechanisms underlying non-relapsing progressive disease, which occurs even in patients diagnosed with RRMS.^17^ The existence of such inflammatory mechanisms is consistent with prior observations that patients on high efficacy DMTs still undergo PIRA,^13,31^ and that while MS DMTs slow brain atrophy (which correlates with PIRA^32,33^), it does not reduce them to levels expected with normal aging.^31^

The lack of significant impact of HEET on risk of PIRA, however, does not detract from data on the importance of timely early HE therapy for preventing overall disability accumulation in MS patients.^3,4^ As demonstrated, HEET significantly reduces risk of RAW, which contributes meaningfully to disability and clinical SPMS conversion.^34,35^ The fact that both RAW and PIRA may contribute to clinical SPMS conversion may also explain some of the differences between this study and the 2019 MSBase study of secondary progressive (SP) MS, which found decreased rates of transition SPMS among patients on fingolimod, alemtuzumab, or natalizumab vs glatimer acetate and interferon beta.^36^

Of note, this study focuses on intensity of early treatment, which was usually sustained over the entire observation period (especially for HEET), but not guaranteed to be. As such, it cannot be directly compared to other studies which compare the effect of specific DMTs on PIRA, for example the OPERA I and II studies showing a slight delay in PIRA in patients receiving ocrelizumab vs IFN-β.^13^ It should be noted, however, that in the OPERA studies, the risk of PIRA is still substantial in the ocrelizumab treated patients, and the observation period was limited to 2 years (compared to 20 years in our study) which may limit assessment of PIRA.

In this analysis, older age at diagnosis was found to be a significant contributor to risk of PIRA, consistent with prior reports,^18–20^ and to risk of sustained EDSS ≥ 5.0. However, even in young patients, where the effect of aging is minimized, disability progression is a fairly common occurrence regardless of intensity of initial therapy. It is admittedly difficult to disentangle the effect of aging on EDSS scoring, and thus PIRA, as it is difficult to confidently isolate MS-related changes with aging-related changes in longitudinal EDSS measurements. Studying PIRA in younger patients, however, may help limit confounding.

In addition, we find in a subset of patients who had baseline 1.5T MRI measurements that baseline brain parenchymal fraction affects risk of PIRA, but not RAW. This is consistent with prior observations in the literature.^32,37^ Unfortunately, due to the small numbers of patients with baseline MRI measurements in this cohort, we could not directly test the potentially confounding effect of different baseline MRI measurements on risk of PIRA by treatment group. In future work, expanding the number of patients with MRI measurements in addition to longitudinal EDSS measurements and treatment data could yield a more robust assessment.

Other limitations of this study include its nature as an observational, single-center study with imperfectly distributed data (especially regarding length of time observed and number of patients on HEET vs. LEET), mostly due to the long history of the CLIMB study with many registered patients diagnosed and treated before most HE therapies became available. The distribution of HE and VHE therapies was also not equally representative of all therapies, consisting mostly of S1P modulators and anti-CD20 therapies. Cladribine in particular was not well represented with only 1 patient on this medicine in the cohort. This is significant given cladribine’s better CNS penetration than other HE/VHE therapies, and recent data suggesting a modestly decreased risk of PIRA on this medication.^38^ The calculation of PIRA in this study was also only based on EDSS score, a broad measure of disability which has inherent limitations (i.e. ambulation focused). While we use this as a first-pass signal for progressive disease, it is not a full measure of disability, disease progression, or existence of progressive MS. Incorporation of 9-hole peg test, MRI data, radiological measurements, or biomarkers could further refine the findings. Calculations of dEDSS/dt were also based on single EDSS measurements, which may be affected by visit variability. But as these scores were analyzed in aggregate, it was felt this variability would be normalized across the population.

While we aim for the results to be as generalizable as possible, the single-center nature of the study, capturing mostly patients in a major metropolitan area with above average access to care and a mostly white demographic, likely limits the generalizability of this study to those patients of a similar background. Additional studies looking at more broadly representative patients are necessary to complement these findings and increase generalizability to other populations.

In all, this study suggests the early use of HE/VHE treatments may not significantly impact a RRMS patient’s long-term risk of silent progression or PIRA leading to a sustained EDSS ≥ 5.0, but significantly reduces disability accumulation from RAW and results in a slightly lower average rate of EDSS increase over time. The benefits of HEET on RAW should be sufficient to still encourage the early use of HE treatments in those with active relapsing disease. However, the minimal effects on PIRA should urge vigilance for silent progression even in those patients on HEET. These findings also have implications for understanding the driving pathophysiological mechanisms in progressive MS, and underscore the need for additional therapies that are effective for preventing PIRA and progressive MS.

## Acknowledgements and Contributions

JJY conceived of the study under the mentorship of TC, and performed the bulk of the cohort data parsing, survival analysis, and statistical analysis. NM helped with discussions on PIRA and organization of cohort data. MP manages and curates the Harvard Multiple Sclerosis Patient Data Base. TC oversees the data collection for the CLIMB study and maintenance of the Harvard Multiple Sclerosis Patient Data Base and the Massachusetts General Hospital Pediatric Multiple Sclerosis database, and provided guidance for the manuscript and edits. The authors give thanks to contributions from Brian Healy for help with the PIRA detection algorithm and statistical guidance, as well as Yuriy Fuksenko for guidance in creating the Python script for parsing the cohort data and integrating this with the PIRA/RAW detection algorithm output.

## Supplemental material

Supplemental material (Supplemental Figures 1-4, Supplemental Tables 1-3) is available in the attached file.

## References

1. Cross A, Riley C. Treatment of Multiple Sclerosis. Continuum (Minneap Minn). 2022;28(4):1025–1051. doi:10.1212/CON.0000000000001170

2. Ontaneda D, Tallantyre EC, Tomáš Kalinčík, Kalincik T, Planchon SM, Evangelou N. Early highly effective versus escalation treatment approaches in relapsing multiple sclerosis. Lancet Neurology. 2019;18(10):973–980. doi:10.1016/s1474-4422(19)30151-6

3. Filippi M, Amato MP, Centonze D, et al. Early use of high-efficacy disease–modifying therapies makes the difference in people with multiple sclerosis: an expert opinion. J Neurol. 2022;269(10):5382–5394. doi:10.1007/s00415-022-11193-w

4. Buron MD, Chalmer TA, Sellebjerg F, et al. Initial high-efficacy disease-modifying therapy in multiple sclerosis. Neurology. 2020;95(8):e1041–e1051. doi:10.1212/WNL.0000000000010135

5. Spelman T, Magyari M, Piehl F, et al. Treatment Escalation vs Immediate Initiation of Highly Effective Treatment for Patients With Relapsing-Remitting Multiple Sclerosis: Data From 2 Different National Strategies. JAMA Neurology. 2021;78(10):1197–1204. doi:10.1001/jamaneurol.2021.2738

6. Selmaj K, Cree BAC, Barnett M, Thompson A, Hartung HP. Multiple sclerosis: time for early treatment with high-efficacy drugs. J Neurol. 2024;271(1):105–115. doi:10.1007/s00415-023-11969-8

7. Katharine Harding, Harding K, Owain Williams, et al. Clinical Outcomes of Escalation vs Early Intensive Disease-Modifying Therapy in Patients With Multiple Sclerosis. JAMA Neurology. 2019;76(5):536–541. doi:10.1001/jamaneurol.2018.4905

8. University of California SFMET, Cree BAC, Hollenbach JA, et al. Silent progression in disease activity–free relapsing multiple sclerosis. Annals of Neurology. 2019;85(5):653–666. doi:10.1002/ana.25463

9. Confavreux C, Vukusic S, Moreau T, Adeleine P. Relapses and Progression of Disability in Multiple Sclerosis. New England Journal of Medicine. 2000;343(20):1430–1438. doi:10.1056/NEJM200011163432001

10. Morgan A, Tallantyre E, Ontaneda D. The benefits and risks of escalation versus early highly effective treatment in patients with multiple sclerosis. Expert review of neurotherapeutics. 2023;23(5):433–444. doi:10.1080/14737175.2023.2208347

11. Ziemssen T, U. Engelmann, Engelmann U, et al. Rationale, design, and methods of a non-interventional study to establish safety, effectiveness, quality of life, cognition, health-related and work capacity data on Alemtuzumab in multiple sclerosis patients in Germany (TREAT-MS). BMC Neurology. 2016;16(1):109–109. doi:10.1186/s12883-016-0629-9

12. Ontaneda D, Tallantyre EC, Raza PC, et al. Determining the effectiveness of early intensive versus escalation approaches for the treatment of relapsing-remitting multiple sclerosis: The DELIVER-MS study protocol. Contemp Clin Trials. 2020;95:106009. doi:10.1016/j.cct.2020.106009

13. Kappos L, Wolinsky JS, Giovannoni G, et al. Contribution of Relapse-Independent Progression vs Relapse-Associated Worsening to Overall Confirmed Disability Accumulation in Typical Relapsing Multiple Sclerosis in a Pooled Analysis of 2 Randomized Clinical Trials. JAMA neurology. 2020;77(9):1132–1140. doi:10.1001/JAMANEUROL.2020.1568

14. Ness NH, Schriefer D, Haase R, Ettle B, Cornelissen C, Ziemssen T. Differentiating societal costs of disability worsening in multiple sclerosis. J Neurol. 2020;267(4):1035–1042. doi:10.1007/s00415-019-09676-4

15. Müller J, Cagol A, Lorscheider J, et al. Harmonizing Definitions for Progression Independent of Relapse Activity in Multiple Sclerosis: A Systematic Review. JAMA Neurology. 2023;80(11):1232–1245. doi:10.1001/jamaneurol.2023.3331

16. Müller J, Sharmin S, Lorscheider J, et al. Standardized Definition of Progression Independent of Relapse Activity (PIRA) in Relapsing-Remitting Multiple Sclerosis. JAMA Neurol. Published online April 14, 2025:e250495. doi:10.1001/jamaneurol.2025.0495

17. Lublin FD, Häring DA, Ganjgahi H, et al. How patients with multiple sclerosis acquire disability. Brain. 2022;145(9):3147–3161. doi:10.1093/brain/awac016

18. Portaccio E, Group on behalf of the IMSRC, Bellinvia A, et al. Progression is independent of relapse activity in early multiple sclerosis: a real-life cohort study. Brain. 2022;145(8):2796–2805. doi:10.1093/BRAIN/AWAC111

19. Iaffaldano P, Portaccio E, Lucisano G, et al. Multiple Sclerosis Progression and Relapse Activity in Children. JAMA Neurol. 2024;81(1):50–58. doi:10.1001/jamaneurol.2023.4455

20. Tur C, Carbonell-Mirabent P, Cobo-Calvo Á, et al. Association of Early Progression Independent of Relapse Activity With Long-term Disability After a First Demyelinating Event in Multiple Sclerosis. JAMA Neurology. 2023;80(2):151–160. doi:10.1001/JAMANEUROL.2022.4655

21. Giovannoni G, Popescu V, Wuerfel J, et al. Smouldering multiple sclerosis: the ‘real MS.’ Therapeutic Advances in Neurological Disorders. 2022;15. doi:10.1177/17562864211066751

22. Thompson AJ, Banwell BL, Barkhof F, et al. Diagnosis of multiple sclerosis: 2017 revisions of the McDonald criteria. Lancet Neurol. 2018;17(2):162–173. doi:10.1016/S1474-4422(17)30470-2

23. Kappos L, Butzkueven H, Wiendl H, et al. Greater sensitivity to multiple sclerosis disability worsening and progression events using a roving versus a fixed reference value in a prospective cohort study. *Multiple Sclerosis (Houndmills, Basingstoke*, England). 2018;24(7):963. doi:10.1177/1352458517709619

24. Chua AS, Egorova S, Anderson MC, et al. Handling changes in MRI acquisition parameters in modeling whole brain lesion volume and atrophy data in multiple sclerosis subjects: Comparison of linear mixed-effect models. Neuroimage Clin. 2015;8:606–610. doi:10.1016/j.nicl.2015.06.009

25. Meier DS, Guttmann CRG, Tummala S, et al. Dual-Sensitivity Multiple Sclerosis Lesion and CSF Segmentation for Multichannel 3T Brain MRI. J Neuroimaging. 2018;28(1):36–47. doi:10.1111/jon.12491

26. Wei X, Warfield SK, Zou KH, et al. Quantitative analysis of MRI signal abnormalities of brain white matter with high reproducibility and accuracy. J Magn Reson Imaging. 2002;15(2):203–209. doi:10.1002/jmri.10053

27. Wei X, Guttmann CRG, Warfield SK, Eliasziw M, Mitchell JR. Has your patient’s multiple sclerosis lesion burden or brain atrophy actually changed? Mult Scler. 2004;10(4):402–406. doi:10.1191/1352458504ms1061oa

28. Kikinis R, Shenton ME, Gerig G, et al. Routine quantitative analysis of brain and cerebrospinal fluid spaces with MR imaging. J Magn Reson Imaging. 1992;2(6):619–629. doi:10.1002/jmri.1880020603

29. Liu L, Meier D, Polgar-Turcsanyi M, Karkocha P, Bakshi R, Guttmann CRG. Multiple sclerosis medical image analysis and information management. J Neuroimaging. 2005;15(4 Suppl):103S–117S. doi:10.1177/1051228405282864

30. Dell’Oglio E, Ceccarelli A, Glanz BI, et al. Quantification of Global Cerebral Atrophy in Multiple Sclerosis from 3T MRI Using SPM: The Role of Misclassification Errors. J Neuroimaging. 2015;25(2):191–199. doi:10.1111/jon.12194

31. Bar-Or A, Thanei GA, Harp C, et al. Blood neurofilament light levels predict non-relapsing progression following anti-CD20 therapy in relapsing and primary progressive multiple sclerosis: findings from the ocrelizumab randomised, double-blind phase 3 clinical trials. EBioMedicine. 2023;93:104662. doi:10.1016/j.ebiom.2023.104662

32. Cagol A, Schaedelin S, Barakovic M, et al. Association of Brain Atrophy With Disease Progression Independent of Relapse Activity in Patients With Relapsing Multiple Sclerosis. JAMA Neurology. 2022;79(7):682–692. doi:10.1001/jamaneurol.2022.1025

33. Nakamura K, Husak S, Bermel R, Cohen J, Ontaneda D. Progression independent of relapses in multiple sclerosis is associated with spinal cord and brain atrophy (P10-3.011). Neurology. 2023;100(17_supplement_2):1742. doi:10.1212/WNL.0000000000202091

34. Scott TF, Diehl D, Elmalik W, Gettings EJ, Hackett C, Schramke CJ. Multiple sclerosis relapses contribute to long-term disability. Acta Neurol Scand. 2019;140(5):336–341. doi:10.1111/ane.13149

35. Novotna M, Paz Soldán MM, Abou Zeid N, et al. Poor early relapse recovery affects onset of progressive disease course in multiple sclerosis. Neurology. 2015;85(8):722–729. doi:10.1212/WNL.0000000000001856

36. Brown JWL, Coles A, Horakova D, et al. Association of Initial Disease-Modifying Therapy With Later Conversion to Secondary Progressive Multiple Sclerosis. JAMA. 2019;321(2):175–187. doi:10.1001/jama.2018.20588

37. Tur C, Carbonell-Mirabent P, Otero-Romero S, et al. The Barcelona baseline risk score to predict long-term prognosis after a first demyelinating event: a prospective observational study. The Lancet Regional Health – Europe. 2025;53. doi:10.1016/j.lanepe.2025.101302

38. Guerra T, Copetti M, Zanetta C, et al. The Italian Multiple Sclerosis Register Experience With Cladribine: Impact on Relapses, PIRA, and Treatment Sequencing Strategies Evaluation. Neurol Neuroimmunol Neuroinflamm. 2025;12(4):e200415. doi:10.1212/NXI.0000000000200415

