## Supplemental Figures 1-4, Supplemental Tables 1-3 for "Effect of Early Treatment Intensity on Progression Independent of Relapse Activity and Disability Accumulation in Multiple Sclerosis"

### Supplementary Figures and Tables

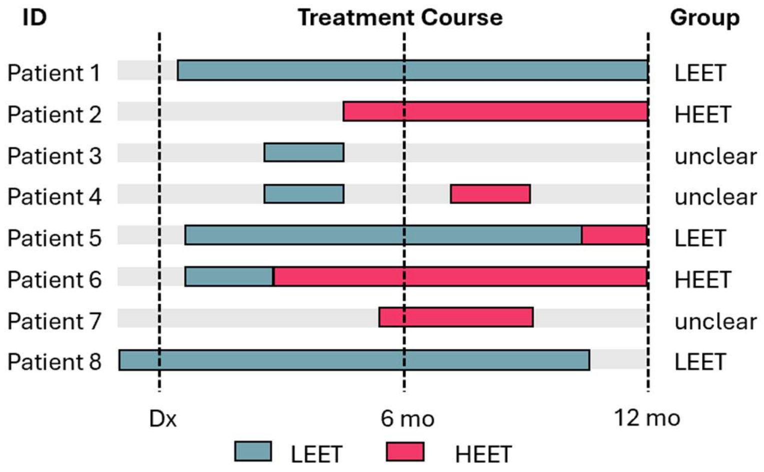

**Supplementary Figure 1. Sorting schema for treatment group based on initial treatment courses.** Examples of various treatment courses and the treatment groups that would be assigned to each patient based on the sorting algorithm (see Methods).

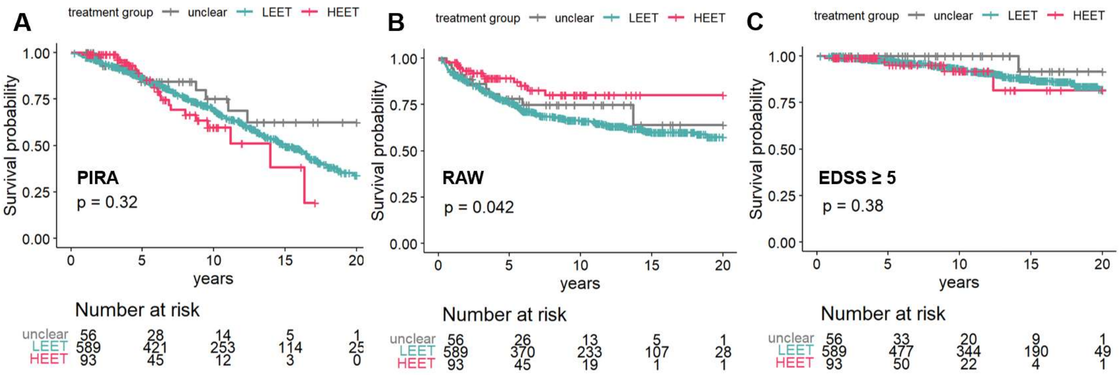

**Supplementary Figure 2.** Event-free survival to first PIRA **(A)**, RAW **(B)**, or sustaining an EDSS ≥ 5.0 **(C)** in all cohort patients, including all MS phenotypes and comparison to patients without a clear initial treatment type (‘unclear’ treatment group). P-values displayed are the result of log-rank tests.

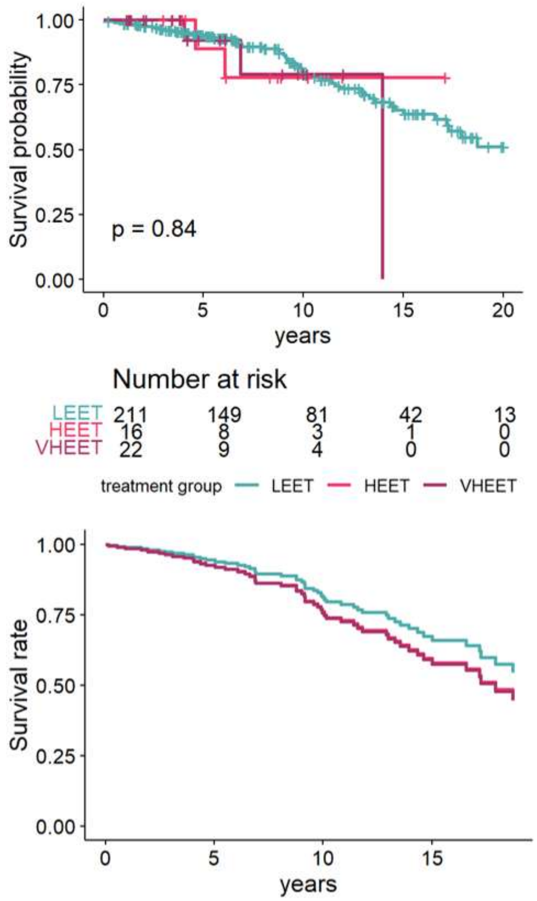

**Supplementary Figure 3.** Event-free survival to first PIRA in young patients (diagnosed with MS before age 30) started on LEET, HEET, or VHEET. Unadjusted survival curves with risk table displayed over adjusted survival curves after fitting a Cox proportional hazards model. P-values displayed are the result of log-rank tests; see **Table 2** for hazard ratios.

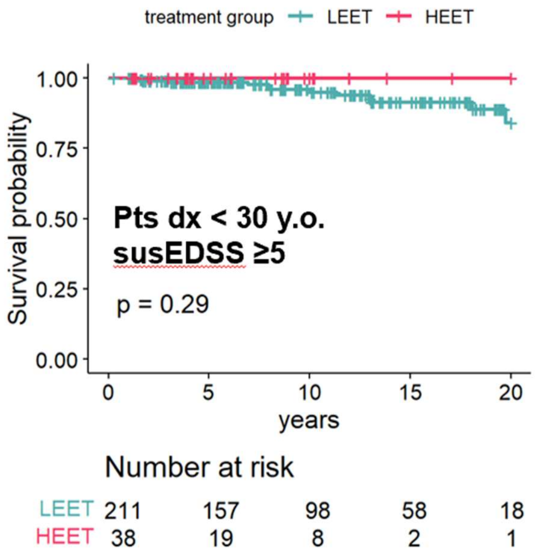

**Supplementary Figure 4.** Event-free survival to sustained EDSS ≥ 5 in young patients (diagnosed at age < 30 years) initiated on LEET vs HEET. P-value displayed is the result of a log-rank test. Hazard ratio unable to be calculated as no events occurred in the HEET group.

**Supplementary Table 1.** Comparison of Patient Characteristics Significantly Different by Treatment Group by ANOVA

| Comparison | Estimate | Std. Error | t value | p-value | |
|---|---|---|---|---|---|
| **first EDSS** | | | | | |
| Low-efficacy treatment - Unclear == 0 | -0.28 | 0.14 | -2.01 | 0.10 | |
| High-efficacy treatment - Unclear == 0 | 0.08 | 0.17 | 0.45 | 0.89 | |
| High-efficacy treatment - Low-efficacy treatment == 0 | 0.36 | 0.11 | 3.25 | 0.003 | ** |
| Low-efficacy treatment - Unclear == 0 | -0.28 | 0.14 | -2.01 | 0.17 | |
| High-efficacy treatment - Unclear == 0 | -0.01 | 0.20 | -0.05 | 1.00 | |
| Very high-efficacy treatment - Unclear == 0 | 0.15 | 0.20 | 0.79 | 0.85 | |
| High-efficacy treatment - Low-efficacy treatment == 0 | 0.27 | 0.15 | 1.79 | 0.26 | |
| Very high-efficacy treatment - Low-efficacy treatment == 0 | 0.44 | 0.15 | 2.95 | 0.02 | * |
| Very high-efficacy treatment - High-efficacy treatment == 0 | 0.16 | 0.21 | 0.80 | 0.84 | |
| **Time in observation** | | | | | |
| Low-efficacy treatment - Unclear == 0 | 4.04 | 0.77 | 5.28 | $<1x10^{-5}$ | *** |
| High-efficacy treatment - Unclear == 0 | -0.92 | 0.92 | -1.00 | 0.57 | |
| High-efficacy treatment - Low-efficacy treatment == 0 | -4.96 | 0.61 | -8.20 | $<1x10^{-5}$ | *** |
| Low-efficacy treatment - Unclear == 0 | 4.04 | 0.77 | 5.27 | $<1x10^{-5}$ | *** |
| High-efficacy treatment - Unclear == 0 | -0.65 | 1.09 | -0.60 | 0.93 | |
| Very high-efficacy treatment - Unclear == 0 | -1.17 | 1.07 | -1.09 | 0.68 | |
| High-efficacy treatment - Low-efficacy treatment == 0 | -4.69 | 0.84 | -5.60 | $<1x10^{-5}$ | *** |
| Very high-efficacy treatment - Low-efficacy treatment == 0 | -5.21 | 0.81 | -6.40 | $<1x10^{-5}$ | *** |
| Very high-efficacy treatment - High-efficacy treatment == 0 | -0.52 | 1.12 | -0.47 | 0.96 | |
| **T2 lesion volume** | | | | | |
| Low-efficacy treatment - Unclear == 0 | 1.26 | 1.22 | 1.03 | 0.55 | |
| High-efficacy treatment - Unclear == 0 | 0.67 | 1.77 | 0.38 | 0.92 | |
| High-efficacy treatment - Low-efficacy treatment == 0 | -0.58 | 1.36 | -0.43 | 0.90 | |
| **BPF** | | | | | |
| Low-efficacy treatment - Unclear == 0 | -1.14 | 1.45 | -0.79 | 0.70 | |
| High-efficacy treatment - Unclear == 0 | -2.66 | 2.10 | -1.27 | 0.41 | |
| High-efficacy treatment - Low-efficacy treatment == 0 | -1.52 | 1.61 | -0.94 | 0.61 | |

Significance codes: 0 ‘***’ 0.001 ‘**’ 0.01 ‘*’ 0.05 ‘.’ 0

**Supplementary Table 2.** Cox Proportional Hazards Output of Treatment Group and Covariates Affecting Risk of PIRA, RAW, or Sustained EDSS≥5.

| Outcome | HR | lower .95 | upper .95 | p-adj | |
|---|---|---|---|---|---|
| **PIRA** (excluding unclear group) (n= 693, events= 250) | | | | | |
| Treatment group - High-efficacy treatment vs reference | 1.27 | 0.81 | 1.98 | 0.31 | |
| Age at diagnosis | 1.03 | 1.02 | 1.05 | $1.32 \times 10^{-5}$ | *** |
| Gender – F vs reference | 0.93 | 0.69 | 1.24 | 0.62 | |
| Race – White vs reference | 1.06 | 0.72 | 1.55 | 0.78 | |
| Initial EDSS | 0.99 | 0.87 | 1.13 | 0.86 | |
| **RAW** (excluding unclear group) (n= 693, events= 208) | | | | | |
| Treatment group - High-efficacy treatment vs reference | 0.51 | 0.29 | 0.89 | 0.02 | * |
| Age at diagnosis | 0.99 | 0.98 | 1.01 | 0.22 | |
| Gender – F vs reference | 1.33 | 0.96 | 1.85 | 0.09 | . |
| Race – White vs reference | 1.04 | 0.70 | 1.55 | 0.85 | |
| Initial EDSS | 1.01 | 0.88 | 1.17 | 0.84 | |
| **susEDSS≥5** (excluding unclear group) (n= 693, events= 69) | | | | | |
| Treatment group - High-efficacy treatment vs reference | 0.91 | 0.36 | 2.33 | 0.85 | |
| Age at diagnosis | 1.05 | 1.02 | 1.09 | $1.06 \times 10^{-3}$ | ** |
| Gender – F vs reference | 0.58 | 0.34 | 1.00 | 0.05 | * |
| Race – White vs reference | 0.66 | 0.33 | 1.34 | 0.25 | |
| Initial EDSS | 2.11 | 1.65 | 2.71 | $4.32 \times 10^{-9}$ | *** |

Significance codes: 0 '***' 0.001 '**' 0.01 '*' 0.05 '.' 0

**Supplementary Table 3.** Cox Proportional Hazards Analysis of Treatment Group, Demographic Covariates, and Baseline MRI Parameters in Patients with 1.5T Baseline MRIs.

| Outcome | HR | lower .95 | upper .95 | p-adj | |
|---|---|---|---|---|---|
| **PIRA** (excluding unclear group) (n= 69, events= 33) | | | | | |
| Treatment group - High-efficacy treatment vs reference | Unable to calculate (no PIRA events in the 4 patients on HEET in this subgroup) | | | | |
| Age at diagnosis (years) | 0.98 | 0.93 | 1.03 | 0.48 | |
| Gender – F vs reference | 0.34 | 0.15 | 0.78 | 0.01 | * |
| Race – White vs reference | 0.95 | 0.32 | 2.88 | 0.93 | |
| Initial EDSS | 0.81 | 0.54 | 1.22 | 0.32 | |
| Baseline T2 lesion volume | 1.07 | 0.92 | 1.24 | 0.37 | |
| Baseline brain parenchymal fraction (%) | 0.80 | 0.70 | 0.91 | $8.09 \times 10^{-4}$ | *** |
| **RAW** (excluding unclear group) (n= 69, events= 34) | | | | | |
| Treatment group - High-efficacy treatment vs reference | 1.36 | 0.30 | 6.13 | 0.69 | |
| Age at diagnosis | 0.97 | 0.93 | 1.02 | 0.22 | |
| Gender – F vs reference | 3.05 | 1.05 | 8.90 | 0.04 | * |
| Race – White vs reference | 1.02 | 0.34 | 3.08 | 0.97 | |
| Initial EDSS | 0.70 | 0.46 | 1.05 | 0.08 | . |
| Baseline T2 lesion volume | 0.94 | 0.81 | 1.09 | 0.41 | |
| Baseline brain parenchymal fraction (%) | 0.92 | 0.81 | 1.05 | 0.20 | |

Significance codes: 0 '***' 0.001 '**' 0.01 '*' 0.05 '.' 0
